# Design of a Secure Wearable Health Data Sharing Platform for Region Hovedstaden: A FHIR DK and GDPR-Compliant Service Architecture

**DOI:** 10.64898/2026.03.12.26348210

**Authors:** Abir Chowdhury, Arshi Irtiza

## Abstract

The 1.8 million residents of Region Hovedstaden (Denmark’s Capital Region) currently lack a secure, standardized pathway for integrating continuous wearable health data into Sundhed.dk, the national electronic health record. Consumer wearables such as Apple Watch, Oura Ring, and Garmin generate longitudinal physiological data relevant to chronic disease management, yet existing workflows rely on manual, non-standardized exports incompatible with FHIR DK v6.0.2 profiles and GDPR Article 25 privacy-by-design requirements. This paper presents a conceptual five-layer microservice architecture for secure wearable data sharing, employing MitID national authentication, National Service Infrastructure (NSI) integration, and Zero Trust security controls. Requirements were derived from a mixed-methods study including surveys of 47 Danish stakeholders and systematic benchmarking of existing platforms. Results show 51.1% conditional willingness to share wearable data under secure conditions, with audit transparency and non-medical misuse identified as central trust factors. Fourteen MoSCoW-prioritized requirements (F1–F7, NF1–NF7) are mapped to architecture components, providing a traceable blueprint for closing the interoperability gap in Danish public healthcare.

## I. INTRODUCTION

The rapid proliferation of consumer-grade wearable devices—including smartwatches, continuous glucose monitors (CGMs), and cardiac sensors—is generating unprecedented volumes of longitudinal physiological data. In Region Hovedstaden, Denmark, chronic disease burden is substantial: diabetes prevalence stands at 7.5% and hypertension affects approximately 28% of the population [1]. Remote patient monitoring studies indicate that integrating such data into clinical workflows can reduce hospital readmissions by approximately 38% [2]. Despite this potential, wearable-generated health data rarely reaches clinicians at hospitals such as Rigshospitalet or Amager Hvidovre Hospital.

Three structural barriers perpetuate this gap. First, Sundhed.dk—the national EHR—lacks dedicated FHIR DK v6.0.2 profiles for continuous patient-generated health data (PGHD) [3]. Second, no authentication pathway enables Danish patients to share wearable data via MitID, leaving Apple HealthKit and Google Fit locked in proprietary ecosystems incompatible with NSI interoperability standards [4]. Third, existing solutions do not satisfy Datatilsynet’s GDPR Article 25 requirements for privacy-by-design, granular consent, and patient-visible audit trails [5].

This paper presents: (1) a MoSCoW-prioritized requirements specification derived from 47-respondent stakeholder surveys and Danish regulatory constraints; and (2) a five-layer conceptual microservice architecture integrating wearable IoT data with Sundhed.dk via FHIR DK, NSI, MitID, and Zero Trust security controls. The contribution is a traceable, standards-aligned design blueprint empirically grounded in Copenhagen stakeholder evidence.

## II. BACKGROUND AND RELATED WORK

### A. IoT in Danish Healthcare

Healthcare IoT systems combine heterogeneous wearable sensors—ECG, PPG, accelerometers—transmitting via BLE or MQTT at high sampling rates [6]. While automated, standards-based pipelines are theoretically feasible, no standardized mechanism currently transforms raw wearable streams into FHIR DK Observation resources for autonomous upload to Sundhed.dk [3]. European PGHD research demonstrates the clinical value of such integration when models are built on interoperable standards and coupled to governance mechanisms for consent and auditability [7].

### B. Danish e-Health Infrastructure

Sundhed.dk provides national EHR access for all Danish citizens but lacks wearable-specific FHIR DK profiles for continuous data streams. The National Service Infrastructure (NSI v3.2), maintained by MedCom, governs messaging and interoperability across Denmark’s 98 municipalities [8]. MitID 2.0 serves as the national identity provider but is absent from all current wearable data pipelines [9]. Datatilsynet—the Danish Data Protection Authority—has issued significant fines for GDPR-non-compliant health data handling, demonstrating that compliance is an operational rather than theoretical concern [5].

### C. Platform Benchmarking

A comparative analysis of four platforms against Danish requirements reveals a persistent gap. Sundhed.dk (7.2/10) lacks wearable FHIR profiles; Apple HealthKit (6.8/10) has no MitID/NSI integration; Google Fit (7.2/10) provides no granular consent; Epic MyChart (8.1/10) offers strong FHIR support but is not aligned with Danish governance assumptions. No platform simultaneously satisfies all four criteria, motivating a dedicated integration layer.

**Table I:** Platform Comparison Against Danish Requirements.

| Platform | FHIR DK | MitID/NSI | Consent/Audit | Score |
| --- | --- | --- | --- | --- |
| Sundhed.dk | Partial | Yes | No | 7.2/10 |
| Apple HealthKit | No | No | No | 6.8/10 |
| Google Fit | No | No | Partial | 7.2/10 |
| Epic MyChart | Partial | No | Partial | 8.1/10 |
| Proposed | Full | Full | Full | — |

## III. METHODOLOGY

This work follows Aalborg University’s Problem-Based Learning (PBL) methodology [10], combining pragmatic research philosophy with three complementary evidence streams.

### Systematic literature review

A 5W search strategy targeted Danish-specific sources including Sundhed.dk/FHIR DK documentation, NSI specifications, Datatilsynet enforcement cases, and MitID integration guidelines.

### Platform benchmarking

Sundhed.dk, Apple HealthKit, Google Fit, and Epic MyChart were scored against four criteria: FHIR DK profile support, MitID/NSI compatibility, consent granularity, and clinical workflow feasibility.

### Stakeholder surveys

Two survey instruments (mini-survey: n=34, September–October 2025; interview-style: n=13 retained from 48 raw submissions after automated-response filtering, December 2025) were distributed via LinkedIn and social media. One expert email validation was collected. The consolidated dataset comprised 47 verified responses. Data quality screening used timestamp clustering and repeated-pattern detection to exclude 35 likely automated submissions.

### Ethics statement

This study was conducted in accordance with Aalborg University’s research ethics guidelines. Survey participation was voluntary and anonymous, with informed consent obtained from all respondents. No sensitive personal data was collected or retained.

Requirements were prioritized using the MoSCoW method per ISO/IEC/IEEE 29148, with each requirement linked to stakeholder evidence, regulatory standards, and architecture components through a three-directional traceability matrix.

## IV. REQUIREMENTS ENGINEERING

### A. Stakeholder Analysis

Five primary stakeholder groups were identified: patients/citizens (data owners requiring MitID login, consent control, and transparency); GPs/hospitals (requiring FHIR-based Sundhed.dk integration); platform operators (requiring scalability and audit readiness); Datatilsynet (requiring demonstrable GDPR Article 25 compliance); and device vendors (requiring stable MQTT/BLE/REST APIs). Survey findings confirmed that citizens treat trust—operationalized as audit visibility and control over data access—as the primary adoption criterion.

### B. Functional Requirements

**Table II:** Functional Requirements (MoSCoW)

| ID | Requirement | Priority |
| --- | --- | --- |
| F1 | Secure MitID/NemLog-in authentication | MUST |
| F2 | Granular, revocable consent (type/provider/time) | MUST |
| F3 | FHIR DK v6.0.2 wearable data import/export | MUST |
| F4 | Real-time audit logging exposed to patients/admins | SHOULD |
| F5 | IoT device integration via MQTT, BLE, REST | SHOULD |
| F6 | Configurable clinician dashboards and alerts | COULD |
| F7 | Blockchain consent receipts | WON'T |

### C. Non-Functional Requirements

**Table III:** Non-Functional Requirements (MoSCoW)

| ID | Category | Target | Priority |
| --- | --- | --- | --- |
| NF1 | Security | TLS 1.3, AES-256, mTLS | MUST |
| NF2 | Performance | <3s API response (P95) | MUST |
| NF3 | Scalability | ≥10,000 concurrent patients | SHOULD |
| NF4 | Availability | ≥99.9% monthly uptime | MUST |
| NF5 | Usability | Onboarding ≤5 min, ≤3 steps | SHOULD |
| NF6 | Compliance | DPIA docs generated <24h | MUST |
| NF7 | Maintainability | CI/CD, ≥80% test coverage | SHOULD |

## V. CONCEPTUAL SERVICE ARCHITECTURE

### A. Architectural Principles

The architecture is governed by five principles: (1) Separation of Concerns via a five-layer model minimizing inter-layer coupling; (2) Standards Compliance with FHIR DK v6.0.2, NSI v3.2, and MitID OIDC flows; (3) Privacy by Design embedding consent enforcement, data minimization, and audit logging as non-negotiable GDPR Article 25 controls; (4) Event-Driven Integration using Apache Kafka to decouple wearable data ingestion from downstream clinical processing; and (5) Zero Trust Security requiring continuous authentication and authorization at all service boundaries using mTLS and token-based controls.

### B. Five-Layer Reference Model

**Table IV:** Five-Layer Architecture Stack.

| Layer | Components | Technologies | Req. |
| --- | --- | --- | --- |
| Device | IoT Gateways, BLE adapters | MQTT 5.0, BLE | F5, NF3 |
| API Gateway | MitID Auth, FHIR API, Consent Middleware | Kong, OAuth 2.1, OIDC | F1, F2, F4, NF2 |
| Business Logic | Consent Service, Access Control, Audit Service | Node.js, Kafka Streams | F2, F4, NF4 |
| Data | FHIR Repository, Audit Logs, Consent Store | MongoDB, PostgreSQL, ClickHouse | F3, NF1, NF6 |
| Presentation | Patient Portal, Clinician Dashboard | React.js, FHIR Viewer | F1, F3, NF5 |

### C. Data Flow Sequence

A representative end-to-end flow: (1) Wearable measurements are collected via BLE and normalized at the Device Layer, producing consistent internal representations with timestamps and device identifiers. (2) Normalized events are published to a Kafka ingestion topic over TLS 1.3. (3) The Consent Service verifies valid, active consent for the data type and recipient context before any processing proceeds—absent consent halts the pipeline and logs the decision. (4) Validated events are transformed into FHIR DK Observation resources and persisted to the FHIR Repository. (5) Clinician access is authenticated via MitID/OIDC at the API Gateway, authorized against current consent scope, and each sensitive operation generates an immutable audit event. (6) Consent revocations are propagated to subsequent ingestion and read decisions, supporting GDPR revocation requirements.

### D. Security and Compliance Controls

The Zero Trust model enforces mTLS across all internal service boundaries and re-checks consent on sensitive read/write operations. The API Gateway applies rate limiting and WAF policies against DoS and API abuse. AES-256 is used at rest and TLS 1.3 in transit. The Reporting Module generates DPIA-ready audit documentation within 24 hours on regulator request, satisfying NF6 and GDPR Article 5(2) accountability requirements.

## VI. SURVEY RESULTS AND REQUIREMENTS VALIDATION

### A. Demographics and Wearable Usage

Of 47 verified respondents, 36.2% reported regular wearable use, 48.9% had never used wearables, and 10.6% used them occasionally. The most commonly tracked metrics were steps/activity (n=24), heart rate (n=16), and sleep (n=12)—directly corresponding to the FHIR DK Observation types targeted by the proposed platform. The high proportion of non-users (48.9%) underscores the importance of the simplified MitID-based onboarding specified in NF5.

### B. Willingness to Share Data

When asked whether they would share wearable data with healthcare professionals under secure conditions, 51.1% responded affirmatively, 38.3% conditionally, and 10.6% declined or were unsure. The conditional group exhibited a strong correlation between willingness and confidence in authentication (F1), consent control (F2), and audit transparency (F4), providing direct empirical validation of these requirements.

### C. Privacy Concerns

**Table V:** Privacy Concerns (n=47, multi-select, 82 total selections)

| Concern | Count | % |
| --- | --- | --- |
| Non-medical use (insurance/employers) | 28 | 59.6% |
| Lack of control over data access | 26 | 55.3% |
| Data privacy and hacking | 18 | 38.3% |
| Device reliability | 12 | 25.5% |
| No concerns | 3 | 6.4% |

Governance concerns dominated over technical ones: 59.6% feared non-medical misuse and 55.3% cited lack of control—validating granular consent (F2) and audit logging (F4) as primary design requirements. These findings reinforce GDPR Article 25 privacy-by-design as a practical adoption prerequisite rather than a compliance formality.

### D. Desired Trust Features and Healthcare Impact

Audit transparency/access logs were the most desired trust feature (38.3%), followed by easy consent control (21.3%) and strong encryption (21.3%), directly validating F4, F2, and NF1 respectively. Concerning healthcare impact, 80.9% expected the platform to improve care quality (36.2% significantly, 44.7% somewhat), aligning with literature reporting 38% readmission reduction through IoT-based remote monitoring [2].

## VII. DISCUSSION

The survey results confirm a structurally important finding: Danish citizens are not opposed to wearable data sharing—they are opposed to sharing without adequate trust mechanisms. This implies that the architectural decisions carrying the greatest adoption impact are not performance or scalability parameters but consent visibility and audit transparency. The proposed platform addresses this through F2 (granular consent), F4 (patient-visible audit logs), and the Zero Trust security model, collectively operationalizing GDPR Article 25 at the infrastructure level.

A key limitation is that the architecture remains conceptual. Non-functional targets (99.9% uptime, <3s P95 response time, 10,000 concurrent patients) are design goals requiring empirical validation through prototyping and load testing. The stakeholder sample does not include clinical cohorts, meaning workflow compatibility findings remain preliminary. Sundhed.dk and FHIR DK profiles may also evolve, introducing integration constraints not captured here.

Future work should prioritize a constrained pilot at Rigshospitalet or Amager Hvidovre Hospital, focusing on a limited set of Observation types (heart rate, activity, sleep) for a defined chronic disease patient group. Such a pilot would validate core consent and audit mechanisms under real operational conditions and provide the performance data needed to confirm or refine the non-functional requirements.

## VIII. CONCLUSION

This paper presented a conceptual five-layer service architecture for secure wearable health data sharing in Region Hovedstaden, addressing a critical interoperability gap in Danish public healthcare. The architecture integrates FHIR DK v6.0.2, NSI v3.2, MitID 2.0, and Zero Trust security controls, grounded in 14 MoSCoW-prioritized requirements derived from systematic stakeholder evidence and regulatory analysis.

Survey data from 47 verified Danish respondents demonstrates that 51.1% of citizens are willing to share wearable data under secure conditions, and 80.9% believe such a platform would improve healthcare quality. Trust—operationalized as audit transparency and granular consent control—emerged as the decisive adoption factor, directly validating the platform’s core design priorities.

The proposed architecture and requirements framework provide a traceable blueprint for pilot planning in Region Hovedstaden and a foundation for future national rollout aligned with Denmark’s Digital Health Strategy 2025–2028. A broader implication is that Danish interoperability profiles would benefit from more explicit standardized support for continuous wearable Observation patterns, reducing reliance on ad hoc integration workarounds across the five health regions.

## Data Availability

All data produced in the present study are available upon reasonable request to the authors. Raw survey data is retained by the corresponding author and can be shared with researchers upon reasonable request, subject to appropriate privacy safeguards.

## ACKNOWLEDGEMENTS

The authors thank all 47 survey respondents and the expert stakeholder who provided email validation for this study. Arshi Irtiza contributed through manuscript proofreading and editorial review.

